# Response-Adapted Bladder Preservation in Muscle-Invasive Bladder Cancer: Results of the Phase II RETAIN-2 Trial and Analysis of ctDNA Dynamics

**DOI:** 10.64898/2026.08.24.26361206

**Authors:** Pooja Ghatalia, Eric A. Ross, Li Zhang, Alexander W. MacFarlane, Matthew R. Zibelman, Fern Anari, Phillip H. Abbosh, Cameron Herberts, William Tester, Patrick J. Mille, Tracy L. Rose, Suzanne Cole, Samantha K. Cheung, Punashi Dutta, Shruti Sharma, Adam C. ElNaggar, Minetta C. Liu, James Ryan Mark, Rosalia Viterbo, Eric M. Horwitz, Mark A. Hallman, Andres F. Correa, Marc C. Smaldone, Robert Uzzo, David YT Chen, Kerry S. Campbell, Alexander Kutikov, Elizabeth R. Plimack, Daniel M. Geynisman

**Author notes:** **Corresponding Author:** Pooja Ghatalia, MD, Fox Chase Cancer Center/Temple Health, 333 Cottman Ave, Philadelphia, PA 19111. **Acknowledgement of research support:** This study was funded by Bristol Myers Squibb. **Prior presentation:** Presented at the 2025 and 2026 American Society of Clinical Oncology Genitourinary Cancers Symposium, San Francisco, California, USA, (Abstract #815 and #LBA632). **Trial registration number:** NCT04506554.

## Abstract

**Purpose:** Response-adapted bladder preservation has emerged as a potential alternative to immediate radical cystectomy for selected patients with muscle-invasive bladder cancer (MIBC), but biomarkers to guide treatment de-escalation are lacking. We report the clinical outcomes of the phase II RETAIN-2 trial together with a retrospective circulating tumor DNA (ctDNA) analysis of the RETAIN-1 and RETAIN-2 studies.

**Patients and Methods:** RETAIN-2 prospectively evaluated neoadjuvant accelerated methotrexate, vinblastine, doxorubicin, and cisplatin (AMVAC) plus nivolumab followed by response-adapted management based on clinical restaging. A retrospective tumor-informed ctDNA analysis evaluated longitudinal ctDNA dynamics and associations with clinical outcomes.

**Results:** Seventy-one evaluable patients were enrolled in RETAIN-2. The trial met its primary endpoint, with a 2-year metastasis-free rate of 77.5% after a median follow-up of 34.7 months. Among 22 patients managed with active surveillance, 15 (68.2%) remained metastasis-free with an intact, non-irradiated bladder and 3 (13.6%) developed metastatic disease. In a sensitivity analysis using time to metastasis, the Kaplan–Meier estimated 2-year metastasis-free probability was 83.7% overall and 85.5% with active surveillance. Retrospective ctDNA analyses were performed in 111 patients from RETAIN-1 and RETAIN-2. Baseline and post-treatment ctDNA positivity were associated with metastatic progression and inferior overall survival. Among patients managed with active surveillance who were ctDNA-negative after treatment, the 2-year Kaplan-Meier estimated metastasis-free probability and overall survival were 91% and 97%, respectively. Plasma ctDNA predicted metastatic progression but not intravesical recurrence.

**Conclusion:** Response-adapted bladder preservation after neoadjuvant AMVAC plus nivolumab achieved encouraging long-term outcomes in selected patients with MIBC. Retrospective ctDNA analyses suggest that plasma ctDNA reflects occult systemic disease rather than bladder-confined recurrence and may refine patient selection for bladder preservation. These findings support prospective evaluation of ctDNA-guided strategies while emphasizing the continued need for bladder-directed surveillance and complementary urinary biomarkers.

## Introduction

Muscle-invasive bladder cancer (MIBC) remains a challenging disease. Although perioperative therapy has evolved with the recent incorporation of enfortumab vedotin plus pembrolizumab, radical cystectomy following cisplatin-based neoadjuvant chemotherapy has long been the standard treatment for eligible patients.^1–3^ Despite improved survival, this approach is associated with substantial morbidity and reduced quality of life, and 30%–50% of patients ultimately develop metastatic recurrence.^2, 4–6^

These limitations have driven interest in response-adapted bladder-preservation strategies that reserve definitive local therapy for patients without an adequate response to neoadjuvant treatment. While active surveillance after clinical complete response (cCR) has shown promise, its broader adoption has been limited by imperfect response assessment, discordance between clinical and pathologic staging, and limited prospective evidence supporting the safety of deferring radical cystectomy.^7–13^

Circulating tumor DNA (ctDNA) has emerged as a promising biomarker of molecular residual disease in urothelial carcinoma.^14–17^ However, its role in guiding bladder-preservation strategies remains undefined, particularly in distinguishing patients at risk for systemic metastatic progression from those at risk for bladder-confined recurrence.

The phase II RETAIN program was developed to evaluate a biomarker-driven, response-adapted approach to bladder preservation. RETAIN-1 demonstrated the feasibility of selective bladder preservation after neoadjuvant accelerated methotrexate, vinblastine, doxorubicin, and cisplatin (AMVAC), but recurrence rates remained substantial.^8^ RETAIN-2 was subsequently designed to improve outcomes through the addition of nivolumab to neoadjuvant AMVAC.^18^

Here, we report the primary clinical outcomes of RETAIN-2 together with an exploratory tumor-informed ctDNA analysis of 111 patients enrolled in RETAIN-1 and RETAIN-2.

We evaluated whether plasma ctDNA reflects systemic metastatic risk or intravesical recurrence and whether ctDNA could identify patients suitable for active surveillance following a cCR.

## Methods

### RETAIN-2 study design

RETAIN-2 was a multicenter, single-arm, phase II, investigator-initiated trial conducted at four academic medical centers (**Figure 1A**). Cisplatin-eligible patients with MIBC received three cycles of AMVAC (with G-CSF support) combined with nivolumab.

**Figure 1.**
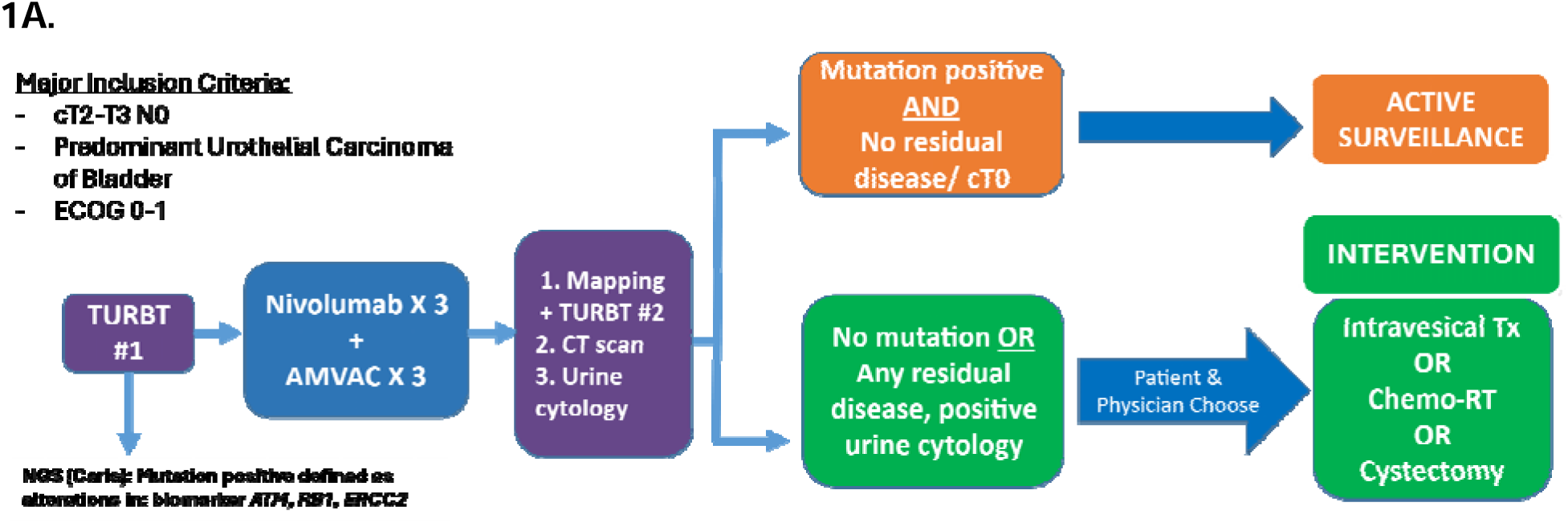
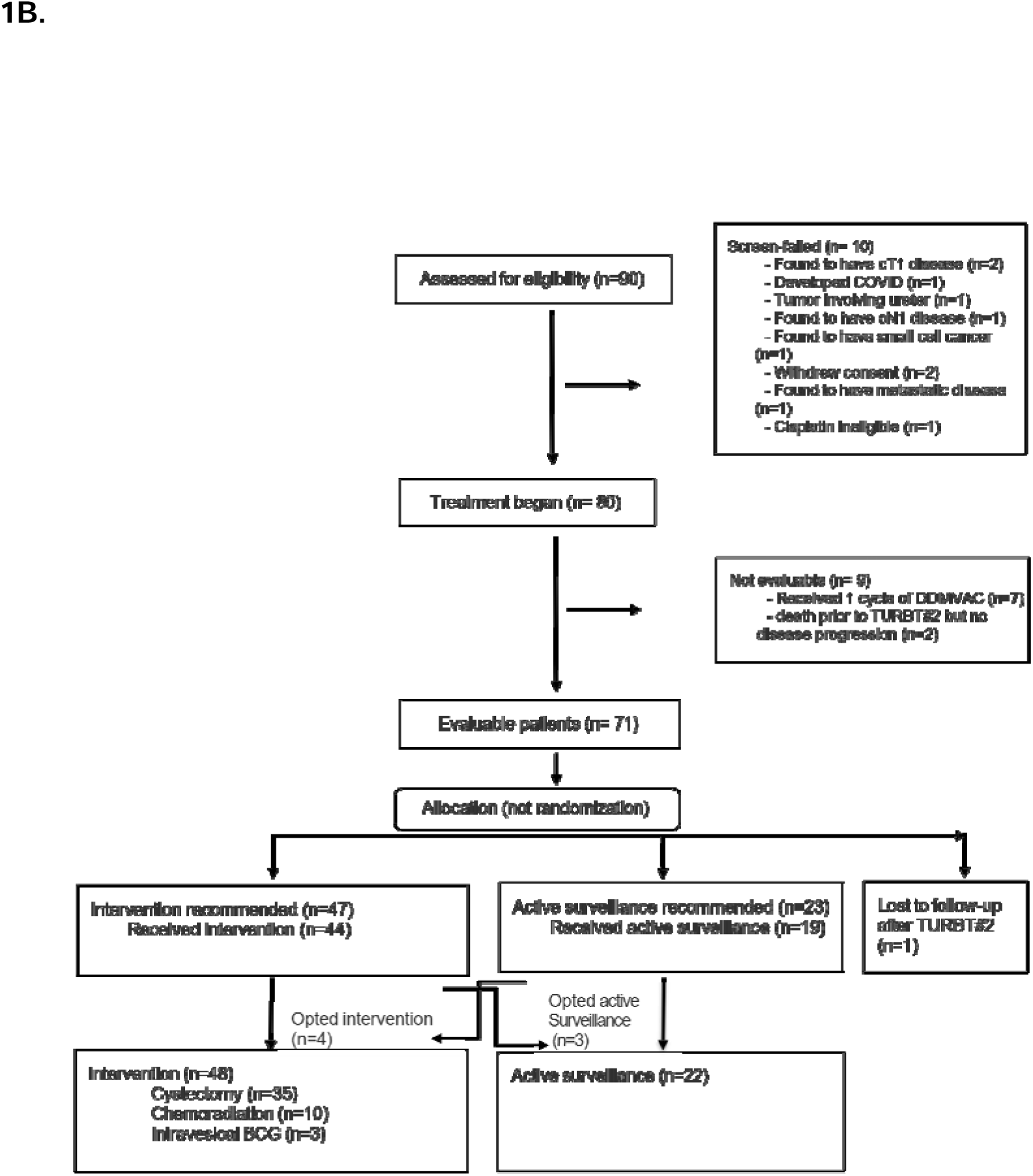
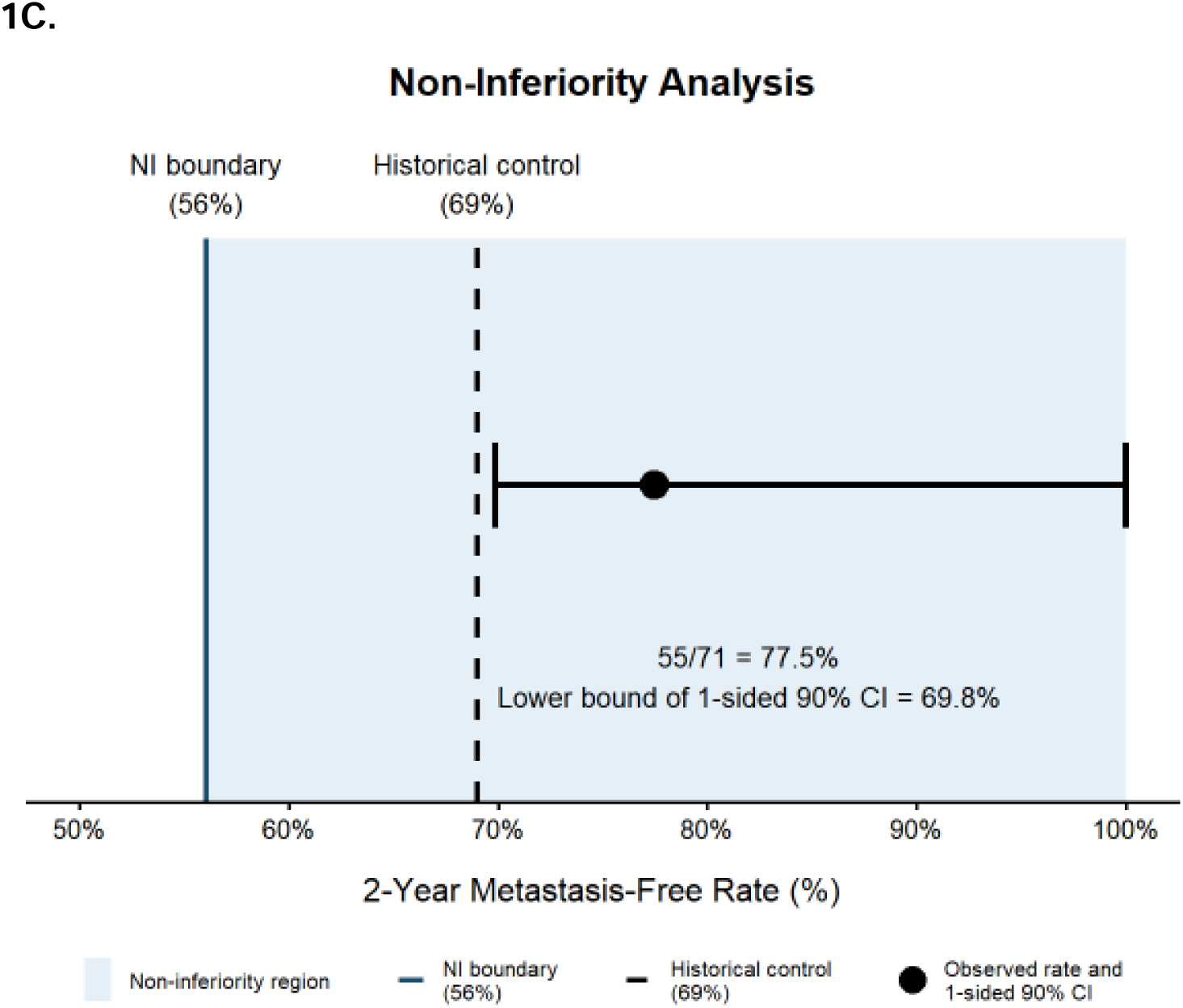
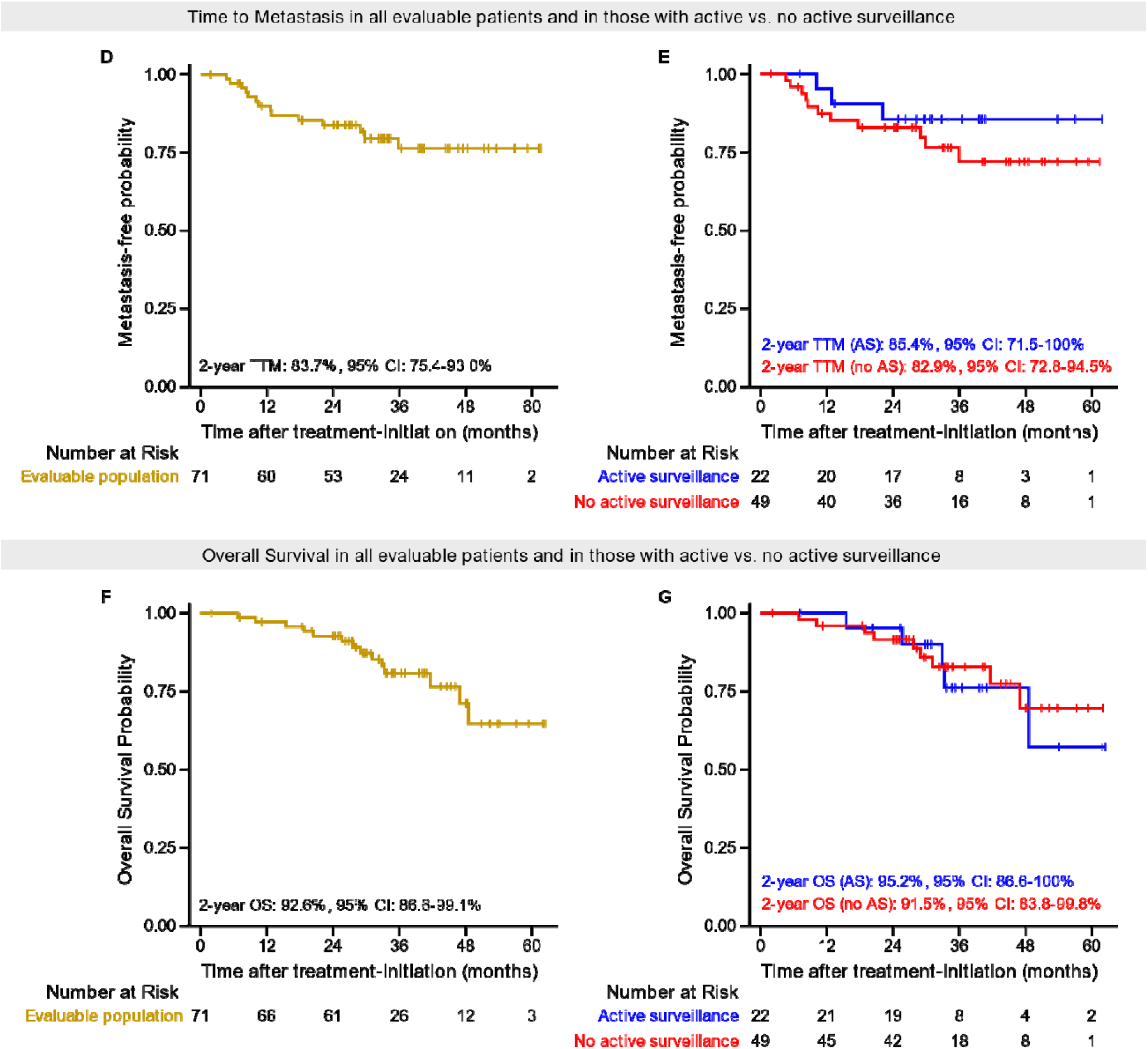
RETAIN-2 trial schema, consort diagram, and clinical outcomes. **A.** Schema of RETAIN-2. **B.** CONSORT diagram of patient enrollment from the RETAIN-2 trial. **C.** Two-year metastasis-free rate in the evaluable population of RETAIN-2. **D.** Kaplan-Meier analysis of time to metastasis in all evaluable patients. **E.** Kaplan-Meier analysis of time to metastasis stratified by treatment group (active surveillance vs intervention). **F.** Kaplan-Meier estimates of overall survival in all evaluable patients **G.** Kaplan-Meier analysis of overall survival stratified by treatment group (active surveillance vs intervention).

Pretreatment transurethral resection of bladder tumor (TURBT#1) specimens underwent molecular profiling to identify alterations in *ATM, RB1*, or *ERCC2*, genomic alterations previously associated with cisplatin response.^19–21^

Following completion of neoadjuvant therapy (NAT), all patients underwent standardized clinical restaging including repeat TURBT (TURBT#2), cross-sectional imaging, and urine cytology. Patient tumors with an *ATM, RB1*, or *ERCC2* mutation who achieved a cCR were offered active surveillance rather than immediate cystectomy or radiation. Patients undergoing surveillance received protocol-defined cystoscopic, cytologic, and radiographic monitoring. Patients who did not meet surveillance criteria were recommended definitive local therapy, including radical cystectomy, chemoradiation, or intravesical therapy, according to protocol and institutional standards.

Patients who completed at least two cycles of both AMVAC and nivolumab were considered evaluable for the primary endpoint. Adjuvant therapy following cystectomy was not prespecified and was administered at investigator’s discretion.

### Patient Eligibility

Eligible patients were ≥18 years old with histologically confirmed urothelial carcinoma, including squamous differentiation or mixed histology, except small cell carcinoma.

Patients with clinical stage T2-T3 N0M0 disease, ECOG performance status 0-1, creatinine clearance ≥ 50 mL/min and adequate bone marrow reserve were eligible. Prior systemic therapy for urothelial carcinoma or cytotoxic chemotherapy for other malignancy within one year was not permitted. This study was registered at ClinicalTrials.gov (NCT04506554), and all patients provided written informed consent. The study was approved by institutional review boards at participating sites and conducted in accordance with the Declaration of Helsinki and Good Clinical Practice guidelines.

### ctDNA analysis across RETAIN-1 and RETAIN-2

Plasma samples from patients enrolled in RETAIN-1 and RETAIN-2 were retrospectively analyzed to evaluate ctDNA associations with clinical outcomes. Both studies enrolled clinically similar populations with MIBC and utilized response-adapted neoadjuvant treatment strategies using neoadjuvant AMVAC alone in RETAIN-1 and AMVAC plus nivolumab in RETAIN-2.^8^ Given shared eligibility criteria, treatment paradigms, response assessments, and follow-up procedures, samples from both trials were analyzed jointly for exploratory biomarker analyses.

ctDNA was assessed using a clinically validated, personalized, tumor-informed, 16-plex mPCR-NGS assay (SignateraTM, Natera, Inc.), as previously described.^22^ Whole-exome sequencing (WES) of pretreatment FFPE tumor tissue and matched normal blood samples. Up to 16 patient-specific somatic single-nucleotide variants (SNVs) were selected for longitudinal plasma-based monitoring. Plasma samples were collected at baseline and after completion of NAT before definitive local therapy in both studies, with additional post-treatment collections at 3 and 6 months in RETAIN-2. ctDNA positivity was defined according to prespecified assay thresholds, and ctDNA levels were reported as mean tumor molecules per milliliter of plasma.

### Immune Profiling by Flow Cytometry

Peripheral blood mononuclear cells (PBMCs) collected from RETAIN-2 participants at baseline and before cycle 2 of treatment were analyzed using multiparameter flow cytometry.^23–25^ Immune profiling evaluated T-cell and natural killer cell subsets, activation markers, proliferation markers, and exhaustion-associated markers. Detailed antibody panels, gating strategies, and analytic methods are provided in the Supplementary Methods.

### Statistical analysis

#### RETAIN-2 clinical endpoints

The primary endpoint was the 2-year metastasis-free rate, defined as the proportion of patients who were free of metastatic urothelial carcinoma 2 years after initiation of NAT. Metastatic progression was defined as lymph node involvement beyond cN1 (more than one clinically suspicious pelvic lymph node), surgically unresectable local disease (e.g., >cT4a), or distant metastasis according to AJCC staging criteria. Patients lost to follow-up before 2 years were considered failures in the primary analysis. Deaths unrelated to urothelial carcinoma were not considered metastatic events.

Key secondary endpoints included any urothelial carcinoma recurrence (including upper tract recurrence) among patients managed with active surveillance, time to metastasis (TTM), overall survival (OS), treatment-related toxicity, and the proportion of patients undergoing radical cystectomy either immediately after NAT or as salvage following surveillance or chemoradiation.

### Study design and Primary Endpoint Analysis

RETAIN-2 was a phase II non-inferiority trial evaluating a risk-adapted bladder-preservation strategy. A 2-year metastasis-free rate of 69% was selected as the efficacy benchmark based on historical outcomes following neoadjuvant cisplatin-based chemotherapy and cystectomy, including approximately 67% recurrence-free survival at 2 years following neoadjuvant AMVAC and cystectomy and similar outcomes reported with neoadjuvant gemcitabine/cisplatin plus pembrolizumab.^27, 28^

The pre-specified non-inferiority margin was 13 percentage points, corresponding to a clinically acceptable lower bound of 56%. This margin was informed by historical 2-year disease-free survival outcomes following neoadjuvant chemotherapy, including 2-year disease-free survival rates of approximately 40% among patients with residual disease at cystectomy, 62% among patients without residual disease after neoadjuvant AMVAC,^29^ and approximately 59% in population-based analyses from the RISC database.^30^

The null hypothesis specified a 2-year metastasis-free rate of 56% and the alternative hypothesis was a rate of 69%. Non-inferiority was assessed using a confidence interval approach. The null hypothesis was rejected if the lower bound of 1-sided 90% exact confidence interval for the observed 2-year metastasis-free rate exceeded 56%.

The planned sample size was 71 evaluable patients. Under the prespecified design, non-inferiority would be demonstrated if at least 46 patients remained metastasis-free at 2 years, corresponding to an observed metastasis-free rate of 64.8%. The design provided approximately 77% power to conclude non-inferiority if the true 2-year metastasis-free rate was 69%, with a one-sided type I error rate of 7.5%. Operating characteristics of the two-stage design were estimated by simulation. The primary endpoint analysis was performed in the protocol-defined evaluable population, consisting of patients who completed at least two cycles of both AMVAC and nivolumab. An exploratory sensitivity analysis evaluated the primary endpoint in the intention-to-treat population, including all enrolled patients regardless of treatment completion.

Prespecified interim monitoring was performed for feasibility and patient safety. The first interim analysis occurred after the first six patients completed 1 year of follow-up, with stopping criteria for excess metastatic events. A second interim review was performed after enrollment of 20 patients with at least 6 months of follow-up, after which the Data and Safety Monitoring Board recommended continuation of the trial.

### Secondary and Exploratory Analyses

As a secondary endpoint, TTM was analyzed using the Kaplan–Meier method. This analysis also served as a sensitivity analysis of the primary endpoint. TTM analysis only considered metastatic progression as an event, and patients without metastatic progression were censored at the date of last disease assessment. Patients lost to follow-up or who died from causes unrelated to urothelial carcinoma were censored at the date of last follow-up or death, respectively.

Exploratory ctDNA analyses evaluated associations between ctDNA status and clinical outcomes. To account for potential immortal time bias in analyses of on-treatment ctDNA, landmark analyses were performed at prespecified sample collection timepoints. Patients experiencing an event before the landmark were excluded, and survival time was recalculated from the landmark date until a subsequent event or censoring.

### General statistical methods

Baseline demographic and clinical characteristics were summarized using descriptive statistics. Treatment-related adverse events were summarized according to CTCAE v5.0 grade.

Time-to-event endpoints, including TTM and OS were estimated using the Kaplan– Meier method. OS was calculated from initiation of NAT until death from any cause, with patients alive at last follow-up censored at their last known date of survival. Hazard ratios (HRs), 95% confidence intervals (CIs), and p-values were estimated using Cox proportional hazards models. Univariable models were used for primary analyses, with multivariable models selectively applied to adjust for potential confounders. Firth-corrected Cox regression was used when model convergence was limited because of low event numbers or collinearity.

All ctDNA analyses were exploratory and hypothesis-generating and results should be interpreted as descriptive, and no adjustment for multiple hypothesis testing was performed unless otherwise specified. Paired immune parameter comparisons were performed using the Wilcoxon signed-rank test, and between-group comparisons used the Wilcoxon rank-sum test.

The primary endpoint was evaluated using a one-sided test consistent with the non-inferiority framework. All other statistical tests were two-sided, with statistical significance defined as P < .05. Analyses were performed using Python and R statistical software.

## Results

### Patient Characteristics and Treatment Delivery

Between December 10, 2020 and April 24, 2024, 90 patients consented and 80 initiated treatment. The data cut-off date for analysis was April 24, 2026, at which time all patients had a minimum of 2 years of follow-up. Nine patients were excluded from the evaluable population, including two who died during NAT and 7 who received only 1 cycle of therapy. The remaining 71 patients comprised the evaluable population (**Table 1**). Sixty patients (75%) completed all three planned cycles of AMVAC plus nivolumab.

Among these evaluable patients, median age was 69 years; 55 (77%) were male, and 66 (93%) were White. Thirty-one patients (44%) harbored a prespecified mutation in *ATM, RB1,* or *ERCC2* and 50 (71%) achieved a cCR following NAT. Based on molecular risk assignment and post-treatment clinical response assessment, 23 patients were assigned to active surveillance and 47 to definitive intervention. One patient was lost to follow-up after TURBT#2 and was not assigned to a treatment pathway (**Figure 1B**).

Ultimately, 22 patients underwent active surveillance and 48 received intervention (cystectomy, N=35; chemoradiation, N=10; intravesical therapy, N=3) (**Figure 1B**).

### Metastasis-free outcomes

At a median follow-up of 34.7 months (interquartile range: 29.5-48.0 months), 55 of 71 evaluable patients remained free of metastatic disease at 2 years, corresponding to a prespecified primary endpoint metastasis-free rate of 77.5%. The lower bound of the one-sided 90% exact confidence interval was 69.8%, exceeding the prespecified non-inferiority boundary of 56% and meeting the primary endpoint criterion (**Figure 1C**).

In an exploratory ITT analysis including all 80 treated patients, the 2-year metastasis-free rate was also 77.5% (62/80). As a sensitivity analysis using a time-to-event framework, the Kaplan-Meier estimated 2-year metastasis-free probability was 83.7% (95% CI, 75.4%-93.0%) in the evaluable population (**Figure 1D**).

In prespecified subgroup analyses, 2-year metastasis-free probabilities were 85.4% (95% CI: 71.5%-100%) in the active surveillance group and 82.9% (95% CI: 72.8%-94.5%) in the intervention group (**Figure 1E**). Two-year OS was 92.6% (95% CI, 86.6%–99.1%) in the evaluable population (**Figure 1F**), and 95.2% (95% CI, 86.6%–100%) and 91.5% (95% CI, 83.8%–99.8%) in the active surveillance and intervention groups, respectively (**Figure 1G**).

Among the 22 patients managed with active surveillance, 9 (41%) developed intravesical recurrence, 1 (4.5%) developed pelvic nodal recurrence and 12 (54.5%) remained free of local or systemic recurrence. Three patients (13.6%) developed metastatic disease, all following prior local recurrence. Four patients underwent salvage cystectomy and three received salvage chemoradiation (**Supplementary Figure 1**).

Final pathology after delayed cystectomy was: ypTaN0 (n=1), ypT2N0 (n=2) and ypT4aN2 (n=2). Overall, 15 of 22 patients (68.2%) remained metastasis-free with an intact, non-irradiated bladder.

Among patients undergoing immediate cystectomy after NAT (N=35), the ypT0 rate was 40% and 63% had <yT2 disease. Eight patients (23%) developed metastatic disease during follow-up. Most ypT0 responses (10/14) occurred among patients without prespecified molecular alterations who underwent cystectomy based on molecular risk assignment rather than residual disease during restaging.

Among patients treated with chemoradiation (n=10), one developed metastatic progression and one died from suspected treatment-related pneumonitis. Among three patients receiving intravesical therapy, one required salvage cystectomy and was found to have ypT4aN0 disease.

### Safety

Grade 3-5 treatment related adverse events (TRAEs) among all 80 treated patients are summarized in **Supplementary Table S1**. Overall, 15 of 80 treated patients (19%) experienced a grade 3-5 TRAE after at least one cycle of AMVAC and nivolumab. Two treatment-related deaths occurred following completion of neoadjuvant therapy, including acute kidney injury and multiorgan failure; both patients were excluded from the evaluable population for the primary endpoint.

### Tumor-Informed ctDNA Predicts Metastatic Risk and Survival

To evaluate the role of ctDNA in response-adapted bladder preservation, plasma samples from RETAIN-1 and RETAIN-2 were analyzed using a personalized tumor-informed ctDNA assay. Both trials enrolled clinically similar patients with MIBC and incorporated response-adapted treatment strategies using neoadjuvant AMVAC alone (RETAIN-1) or AMVAC plus nivolumab (RETAIN-2) (**Supplementary Table S2**). Across both studies, 275 plasma samples from 111 patients (RETAIN-1, n=45; RETAIN-2, n=66) were analyzed (**Supplementary Figure 2**).

Baseline ctDNA was detectable in 42.7% of patients (47/110) and was associated with increased risk of metastatic progression (TTM: HR=7.62, 95% CI: 2.85–20.37; p=0.0001) and death (OS: HR=6.8, 95% CI: 2.27–20.36; p=0.0006) compared with ctDNA-negative patients (**Figure 2A, B**).

**Figure 2.**
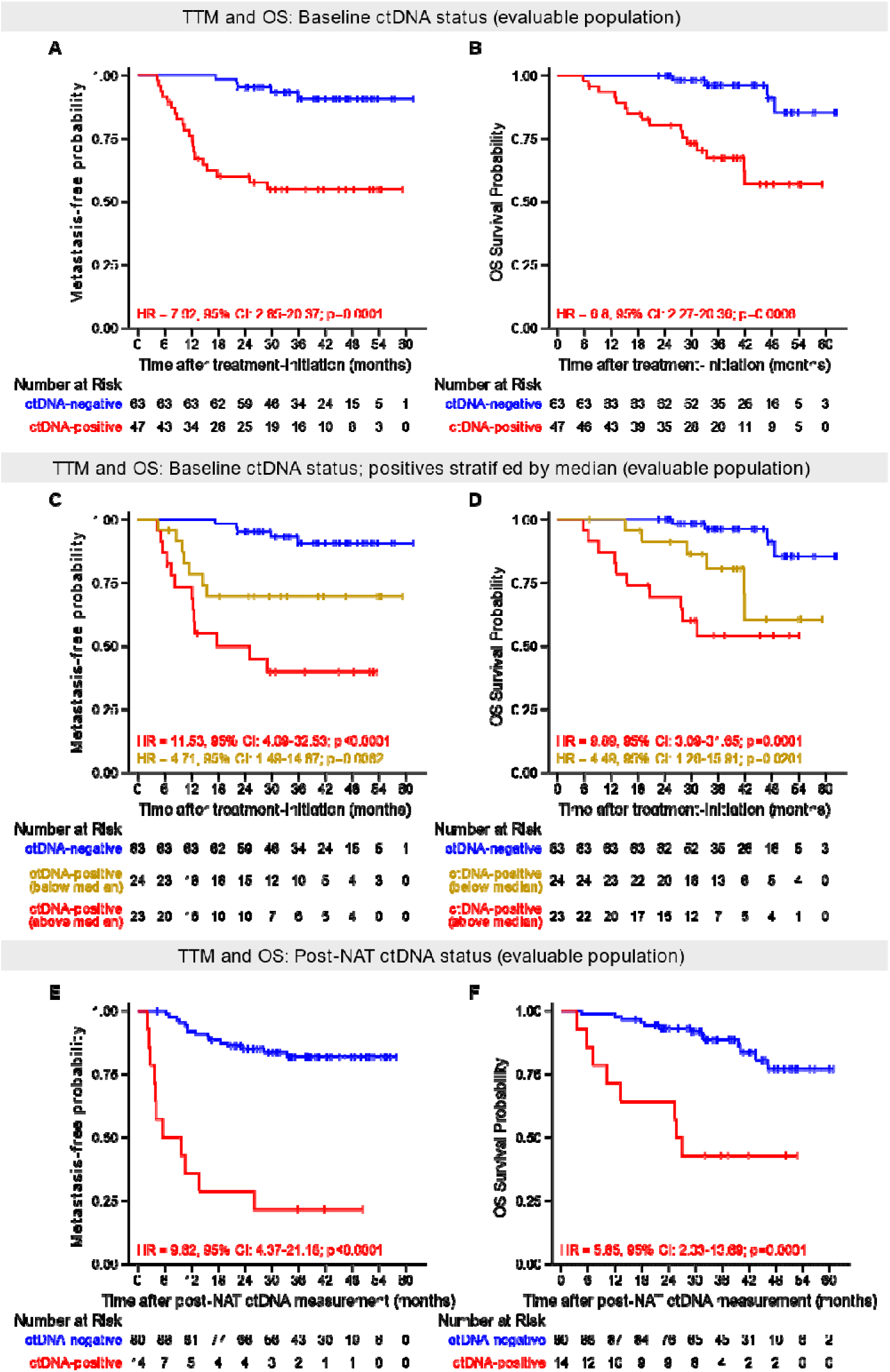
TTM and OS stratified by ctDNA features at baseline and post-NAT in the RETAIN-1 and RETAIN-2 evaluable population. Association of ctDNA status with TTM **(A,C,E)** and OS **(B,D,F).** Kaplan–Meier estimates comparing ctDNA-positive versus ctDNA-negative patients at baseline **(A,B)**, baseline ctDNA stratified by median level **(C,D)**, and post-NAT **(E,F)** timepoints. HRs and 95% CIs were calculated using the Cox proportional hazard models.

Higher baseline ctDNA levels were associated with worse outcomes. Patients with ctDNA levels above the cohort median had the highest risk of metastatic progression (HR=11.53, 95% CI: 4.09–32.53; p<0.0001) and death (HR=9.89, 95% CI: 3.09–31.65; p=0.0001), while patients with detectable ctDNA below the median had intermediate risk outcomes (**Figure 2C, D**).

Following NAT, ctDNA positivity was less common (13.6%; 14/103) but remained strongly prognostic for metastatic progression (HR=9.62, 95% CI: 4.37–21.18; p<0.0001) and death (OS: HR=5.65, 95% CI: 2.33–13.69; p=0.0001) (**Figure 2E, F**). In multivariable analysis incorporating baseline ctDNA, post-NAT ctDNA and post-NAT clinical response, ctDNA positivity at each timepoints remained independently associated with TTM (**Supplementary Table S3**).

Cohort-specific analyses and models incorporating trial cohort as a covariate demonstrated consistent associations between ctDNA status and clinical outcomes, supporting the prognostic performance of ctDNA across both RETAIN-1 and RETAIN-2 (**Supplementary Figure 3; Supplementary Table S4**).

### ctDNA Dynamics During Therapy Identify Distinct Risk Groups

Among patients with detectable baseline ctDNA, 42 of 44 (95.5%) showed ctDNA reduction after NAT and 32 (72.7%) achieved complete ctDNA clearance. Clearance was more common among patients with lower baseline ctDNA levels (median 0.46 vs. 5.61 MTM/mL; p=0.0029). Clearance rates were observed in both patients undergoing cystectomy (61.9%) and those managed with active surveillance (83.3%) (**Figure 3A**).

**Figure 3.**
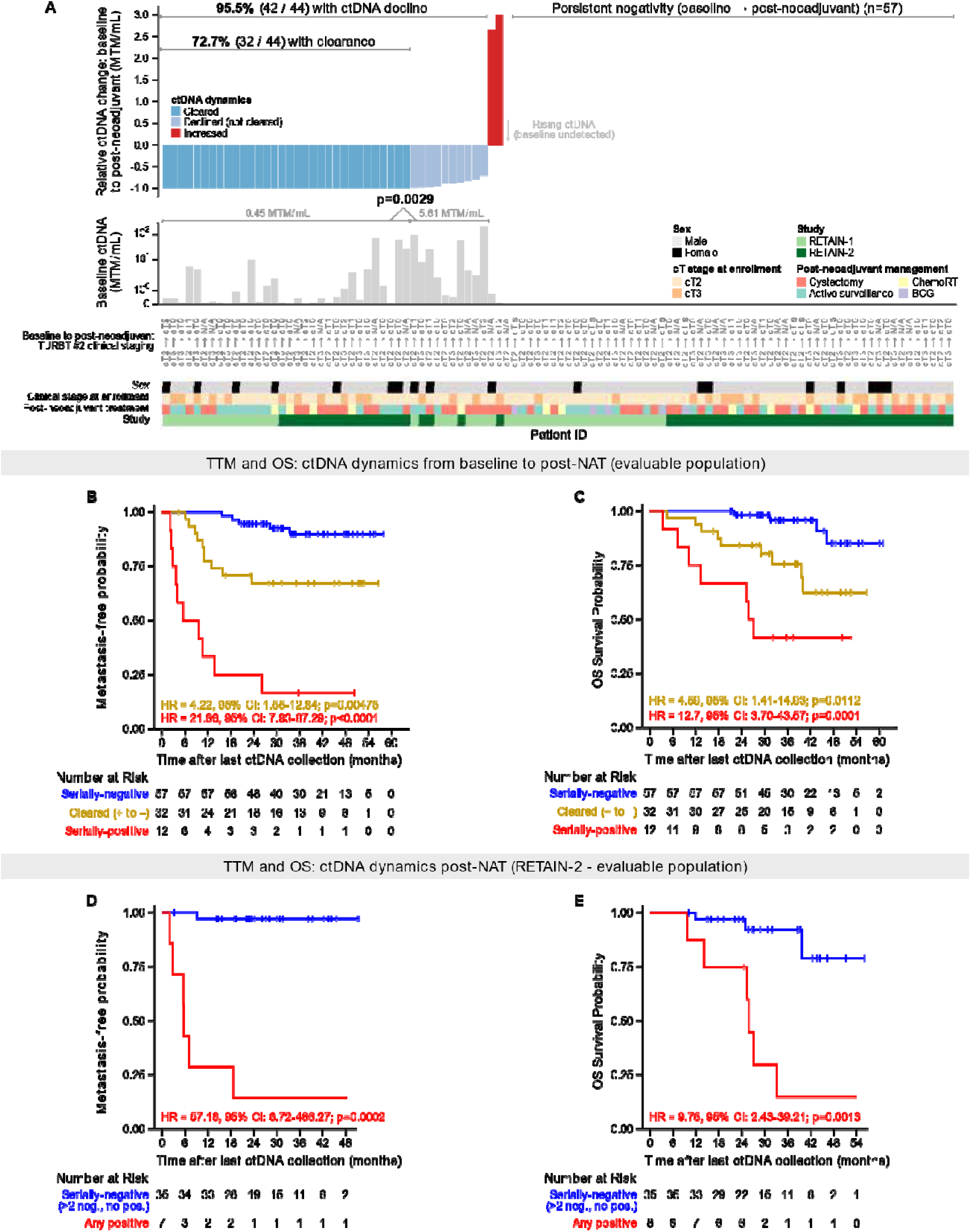
Association of ctDNA dynamics with outcomes. **A.** Patient-level ctDNA dynamics during NAT. Top row: Composite bar plot depicting longitudinal ctDNA dynamics for individual patients (each column represents one patient). Relative change in ctDNA levels (MTM/mL) from baseline (pretreatment) to post-NAT, where -1 (y-axis) denotes complete ctDNA clearance. Relative ctDNA change is only shown for patients with ctDNA positivity at baseline; patients with persistent ctDNA negativity are shown separately (n=57). Middle row: Baseline quantitative ctDNA levels, illustrating lower baseline ctDNA burden among patients who subsequently achieved ctDNA clearance after NAT (Mann-Whitney U test). Bottom rows include annotation of clinical stage change during NAT, post-NAT management approach, and study cohort. Association of ctDNA dynamics from baseline to post-NAT with TTM **(B)** and OS **(C)** Association of longitudinal ctDNA dynamics post-NAT in RETAIN-2 with TTM **(D)** and OS **(E).** For (D) and (E), Kaplan-Meier analyses were performed using a landmark approach at the final ctDNA collection (patients contributed variable numbers of serial timepoints). HRs and 95% CIs were calculated using the Cox proportional hazard models.

Longitudinal ctDNA dynamics were strongly associated with outcomes (**Figure 3B, C**). Patients who remained ctDNA-negative before and after NAT had the most favorable prognosis, with 24- and 36-month metastasis-free probabilities of 94.7% and 89.9%, respectively. In contrast, persistent ctDNA positivity was associated with markedly inferior outcomes (median TTM 5.59 months; median OS 25.79 months) and significantly increased risks of metastatic progression (HR=21.86, 95% CI 7.93–67.29; p<0.0001) and death (HR=12.7, 95% CI 3.70–43.57; p=0.0001) relative to serially negative patients.

Patients who cleared ctDNA during therapy had intermediate outcomes, with higher risks of metastatic progression and death than serially negative patients (TTM: HR=4.22, 95% CI: 1.55–12.84; p=0.0048; OS: HR= 4.59, 95% CI: 1.41–14.93; p=0.011).

Among RETAIN-2 patients with additional ctDNA assessments at 3 and 6 months after NAT, persistent or recurrent ctDNA positivity remained strongly associated with inferior outcomes compared with sustained ctDNA negativity (TTM: HR=57.18, p=0.0002; OS: HR=9.76; p=0.0013) (**Figure 3D, E**).

### Post-NAT ctDNA Stratifies Systemic Risk but Does Not Replace Local Disease Assessment

Among patients who underwent cystectomy, post-NAT ctDNA positivity identified a subgroup at markedly elevated risk of metastatic progression (HR=13.42, 95% CI 4.70– 38.35; p<0.0001) and death (HR=4.56, 95% CI 1.57–13.26; p=0.0053), with a median TTM of only 3.98 months (**Figure 4A, B**).

**Figure 4.**
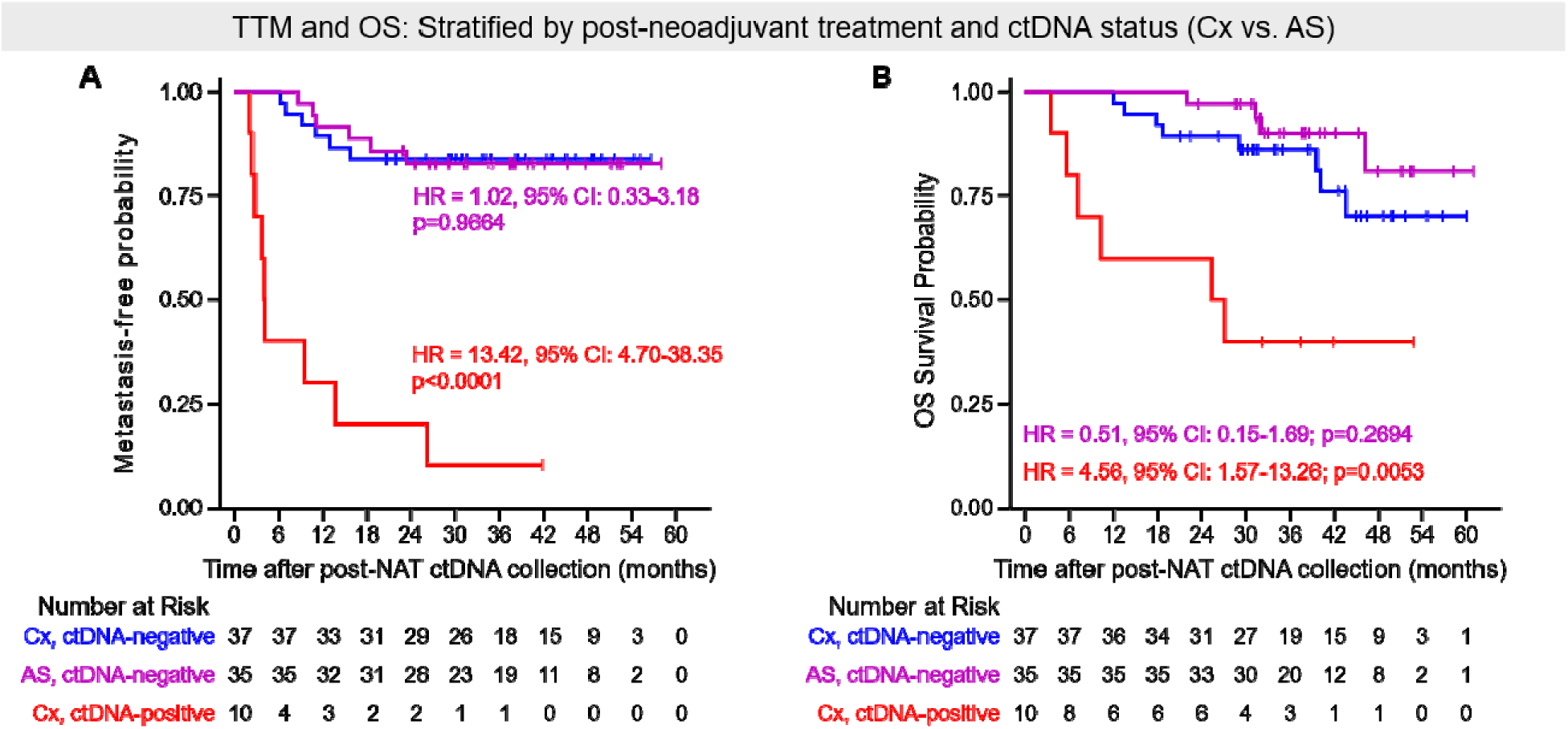
Association of ctDNA status with TTM survival and OS in cystectomy and AS population. Association of ctDNA status with TTM **(A)** and OS **(B).** Kaplan– Meier estimates comparing ctDNA-positive versus ctDNA-negative patients at the post-NAT timepoint. HRs and 95% CIs were calculated using the Cox proportional hazard models.

Among patients who were ctDNA-negative after NAT, outcomes were favorable regardless of whether patients underwent active surveillance or cystectomy, although these non-randomized groups cannot be directly compared. The 24-month metastasis-free probability was 91.4% with active surveillance and 83.8% with cystectomy, while 24-month OS estimates were 97.1% and 89.2%, respectively.

Post-NAT ctDNA also provided prognostic information beyond clinical and pathologic response. Among patients undergoing restaging, ctDNA positivity was associated with increased metastatic risk in both patients with favorable post-NAT bladder findings (cT0/cTis/cTa; HR, 10.86; 95% CI, 2.62-36.37; P=.0025) and those with residual disease (≥cT1; HR, 5.91; 95% CI, 1.55-22.56; P=.0093) (**Figure 5A-B**).

**Figure 5.**
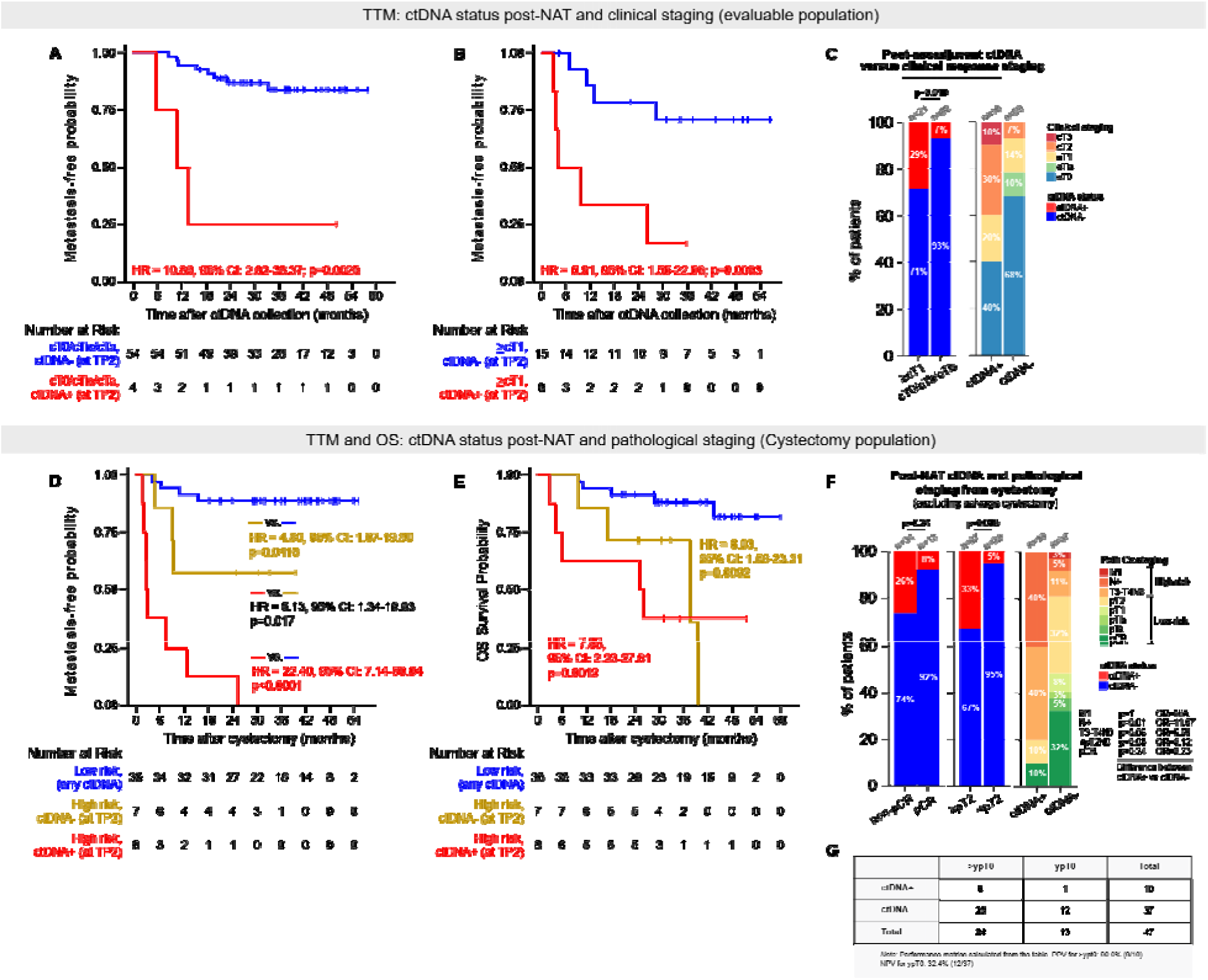
Association of ctDNA status with TTM and OS stratified by clinical and pathological staging. Kaplan–Meier estimates of TTM for ctDNA-positive vs. ctDNA-negative patients across clinical staging groups in the RETAIN-1 and RETAIN-2 evaluable population **(A,B)**, Kaplan–Meier estimates of TTM (**D)** and OS **(E)** for ctDNA-positive versus ctDNA-negative patients stratified by pathologic staging in patients undergoing cystectomy **(D,E)**, Association of ctDNA status with clinical and pathological outcomes. **(C,F)**. **(G)** Positive predictive value (PPV) for residual disease and negative predictive value (NPV) of ctDNA for ypT0 in patients undergoing cystectomy. HRs and 95% CIs were calculated using the Cox proportional hazard models.

Among cystectomy patients, the combination of ctDNA positivity and high-risk pathologic features identified patients with the poorest outcomes. Compared with patients with low-risk pathology (pCR or <pT2N0), patients with ctDNA positivity and high-risk pathologic disease had increased risk of metastatic progression (HR, 22.39; 95% CI, 7.14-80.04; P<.0001) and death (HR, 7.88; 95% CI, 2.23-27.81; P=.0013) (**Figure 5D-E**).

However, ctDNA negativity did not exclude residual bladder disease. Although ctDNA positivity was enriched among patients with residual high-risk disease, 67% of patients with >ypT2 disease were ctDNA-negative, and 51% of ctDNA-negative patients had residual muscle-invasive disease at cystectomy (**Figure 5F**). The positive predictive value of post-NAT ctDNA positivity for residual disease was high (90%), whereas the negative predictive value for ypT0 disease was limited (32.4%), indicating that ctDNA cannot reliably replace local bladder assessment.

### ctDNA Predicts Metastatic Risk but Not Intravesical Recurrence During Active Surveillance

Among patients managed with active surveillance across RETAIN-1 and RETAIN-2, 37 of 40 patients (93%) were ctDNA-negative after treatment and had favorable systemic outcomes, with 24-month metastasis-free probability of 91.4% and overall survival of 97.1%.

However, post-treatment ctDNA status was not predictive of bladder-confined recurrence. Despite negative post-treatment ctDNA, 19 of 37 patients (51%) developed intravesical recurrence during surveillance (**Supplementary Figure 2**). Among patients who developed intravesical recurrence, 19 of 21 (90%) were ctDNA-negative after treatment. These findings indicate that ctDNA is a strong marker of systemic relapse risk but has limited sensitivity for bladder-confined disease.

Exploratory immune profiling was performed in RETAIN-2 participants to evaluate peripheral immune correlates of response. Associations between treatment-induced immune changes and clinical outcomes were observed and are described in the Supplementary Appendix.

## Discussion

RETAIN-2 met its primary endpoint, demonstrating that a response-adapted bladder-preservation strategy incorporating neoadjuvant AMVAC plus nivolumab achieved a 2- year metastasis-free rate consistent with contemporary benchmarks for muscle-invasive bladder cancer (MIBC). Among patients managed with active surveillance, 68% remained metastasis-free with an intact, non-irradiated bladder, supporting the feasibility of selective bladder preservation in appropriately selected patients while avoiding the morbidity associated with immediate radical cystectomy.

Compared with RETAIN-1, RETAIN-2 demonstrated favorable outcomes among patients managed with active surveillance, including lower intravesical recurrence rates (41% vs 62%) and a higher proportion of patients remaining metastasis-free with an intact, non-irradiated bladder (68% vs 48%). Although cross-trial comparisons should be interpreted cautiously, these findings are consistent with emerging evidence supporting perioperative immune checkpoint inhibition in MIBC and suggest that incorporation of nivolumab may enhance systemic disease control within response-adapted treatment strategies.^1, 3, 7^

The genomic selection strategy used in RETAIN-1 and RETAIN-2, based on alterations in *ATM, ERCC2,* and *RB1*, did not consistently identify patients most likely to benefit from bladder preservation. A substantial proportion of patients without qualifying genomic alterations achieved favorable responses, including ypT0 at cystectomy.

Conversely, use of this genomic framework excluded some patients who may have been candidates for bladder preservation. This strategy likely contributed to the high ypT0 rate among patients undergoing cystectomy. Future response-adapted bladder-preservation strategies should prioritize direct assessment of treatment response rather than reliance on limited pretreatment genomic biomarkers.

This study provides one of the largest evaluations of ctDNA within a bladder-preservation framework. Prior studies, including IMvigor011, NIAGARA, INDIBLADE, Epstein et al., SunRISe-4, and HCRN GU16-257, established the prognostic importance of ctDNA in perioperative and bladder-preservation settings; however, most evaluated ctDNA in the context of definitive local therapy.^31–35^ The present analysis extends these findings by evaluating longitudinal ctDNA dynamics among patients managed with and without immediate cystectomy or chemoradiation.

Both baseline and post-NAT ctDNA positivity were strongly associated with metastatic progression and mortality, with consistent findings across multiple analyses. Persistent ctDNA positivity identified a subgroup with markedly increased risk of metastatic progression despite local therapy, suggesting that that ctDNA may help identify patients unlikely to benefit from active surveillance alone. Conversely, patients who remained ctDNA-negative throughout treatment experienced excellent outcomes, supporting the potential role of ctDNA as a biomarker for systemic risk stratification.

Among patients managed with active surveillance who were ctDNA-negative following NAT, 24-month metastasis-free probabilities and OS were 91% and 97%, respectively. Although these findings require prospective validation, they support the hypothesis that ctDNA may complement clinical response assessment to identify patients who can safely defer immediate radical cystectomy after achieving a cCR. In contrast, patients with post-NAT ctDNA positivity had poor outcomes even when treated with local therapy, highlighting the need for additional systemic treatment strategies for this high-risk population.

Importantly, ctDNA appears to reflect systemic residual disease risk rather than residual bladder tumor burden. Although ctDNA positivity was enriched among patients with advanced pathologic disease, ctDNA negativity did not exclude persistent bladder tumor. Therefore, plasma ctDNA should not replace local bladder assessment with cystoscopy, TURBT, or imaging in response-adapted treatment approaches. Complementary biomarkers, including urinary tumor DNA assays, may be required to improve detection of residual intravesical disease.

Several limitations should be considered. First, the primary endpoint was evaluated in a protocol-defined evaluable population, although similar results were observed in the intention-to-treat analysis. Second, the single-arm design requires comparison with historical benchmarks and may be influenced by patient selection, staging accuracy, and institutional expertise. Third, the study was conducted at academic centers, which may limit generalizability. Fourth, longer follow-up is needed to evaluate durability of bladder preservation and late metastatic events. The ctDNA analyses were exploratory and limited by sample size for subgroup analyses and incomplete longitudinal sampling beyond the early post-NAT period. Finally, although exploratory immune profiling suggested potential associations between treatment-induced immune activation and clinical response, these findings require validation in larger prospective cohorts and should be considered hypothesis-generating.

In conclusion, RETAIN-2 demonstrates the feasibility of a response-adapted bladder-preservation strategy incorporating neoadjuvant AMVAC plus nivolumab. Integrated ctDNA analysis suggests that plasma ctDNA provides clinically meaningful information regarding systemic metastatic risk but does not reliably identify residual bladder disease. These findings support prospective evaluation of ctDNA-guided bladder-preservation strategies, particularly as novel perioperative approaches, including enfortumab vedotin plus pembrolizumab, continue to reshape treatment paradigms for MIBC.

## Data Availability

To minimize the risk of patient re-identification, de-identified patient-level clinical data will be made available upon scientifically sound request for academic use only, within the limits of patient informed consent and applicable data protection regulations. Requests should be directed to the corresponding author and will be reviewed by Fox Chase Cancer Center; approved access will require completion of applicable institutional review and a data access agreement. Requests will be answered within 4 weeks.

## ACKNOWLEDGMENTS

We thank the patients who participated in this study, their families, and the investigators and staff at the four study sites, as well as the members of the safety monitoring committee.

