## Supplementary Tables and Figures for "Response-Adapted Bladder Preservation in Muscle-Invasive Bladder Cancer: Results of the Phase II RETAIN-2 Trial and Analysis of ctDNA Dynamics"

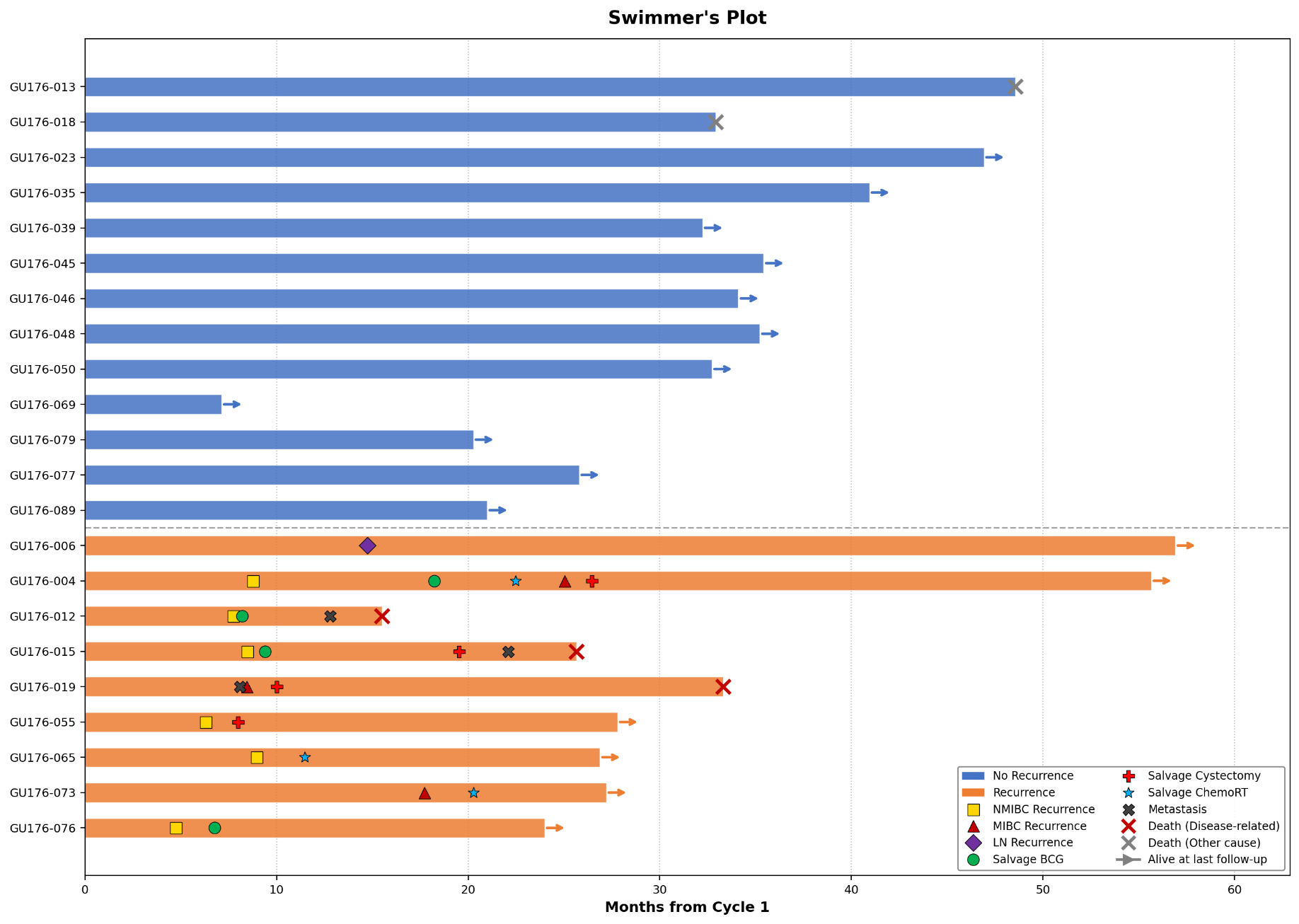

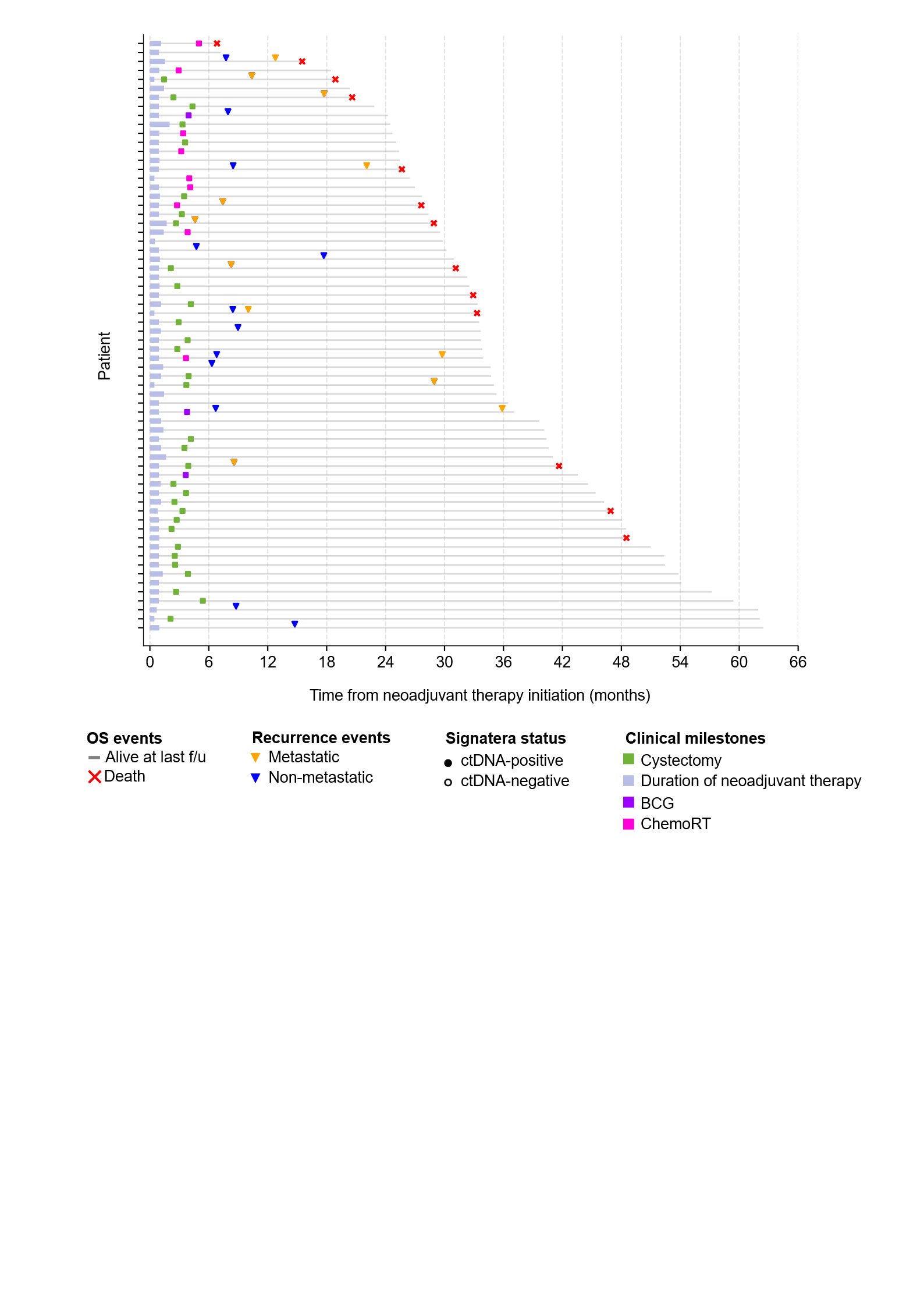

**Supplementary Figure 1: Swimmer’s plot with each lane representing a patient on active surveillance in RETAIN-2 trial**

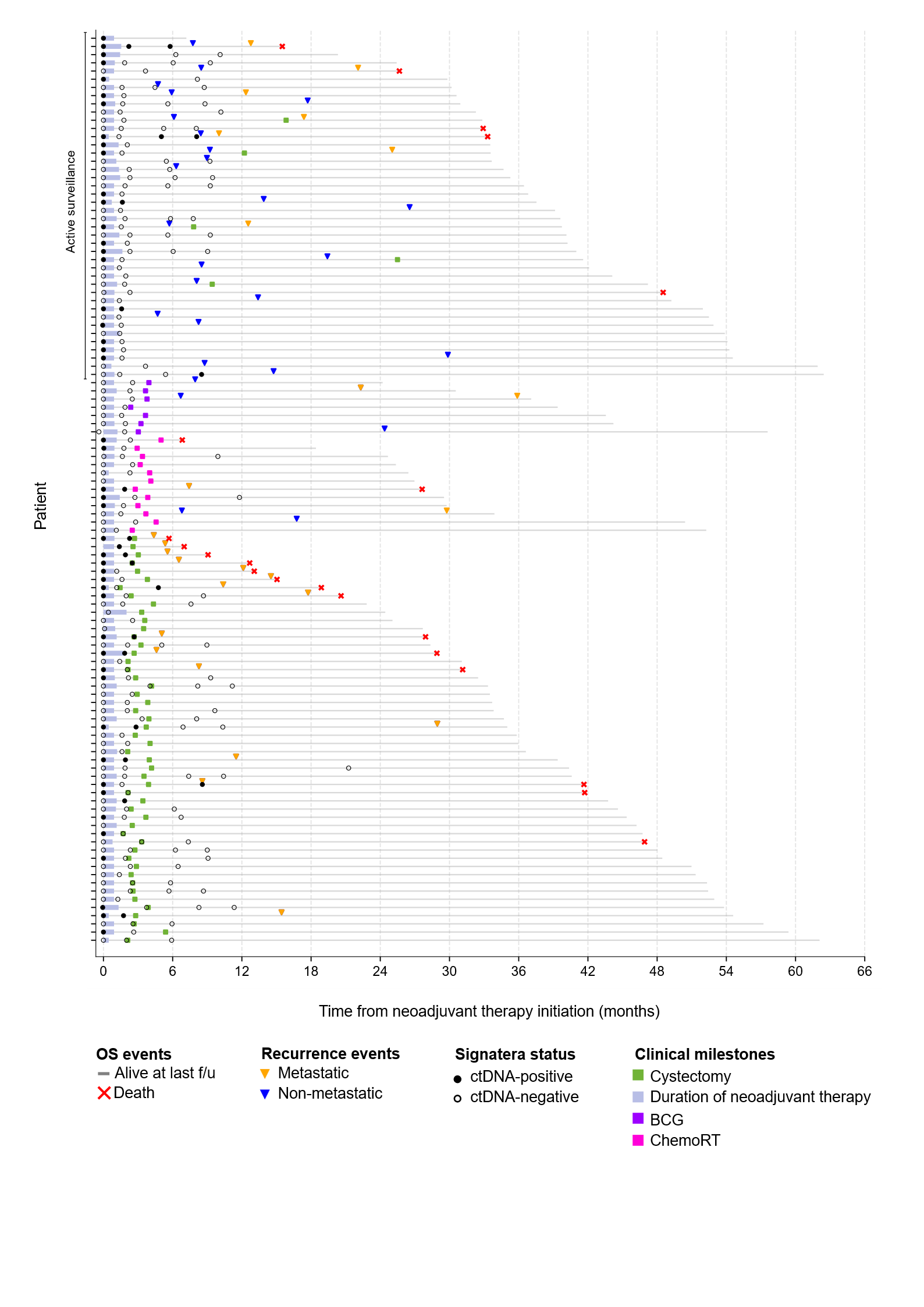
**Supplementary Figure 2: Swimmer’s plot with each lane representing a patient in the integrated ctDNA analysis cohort (RETAIN-1 and RETAIN-2)**

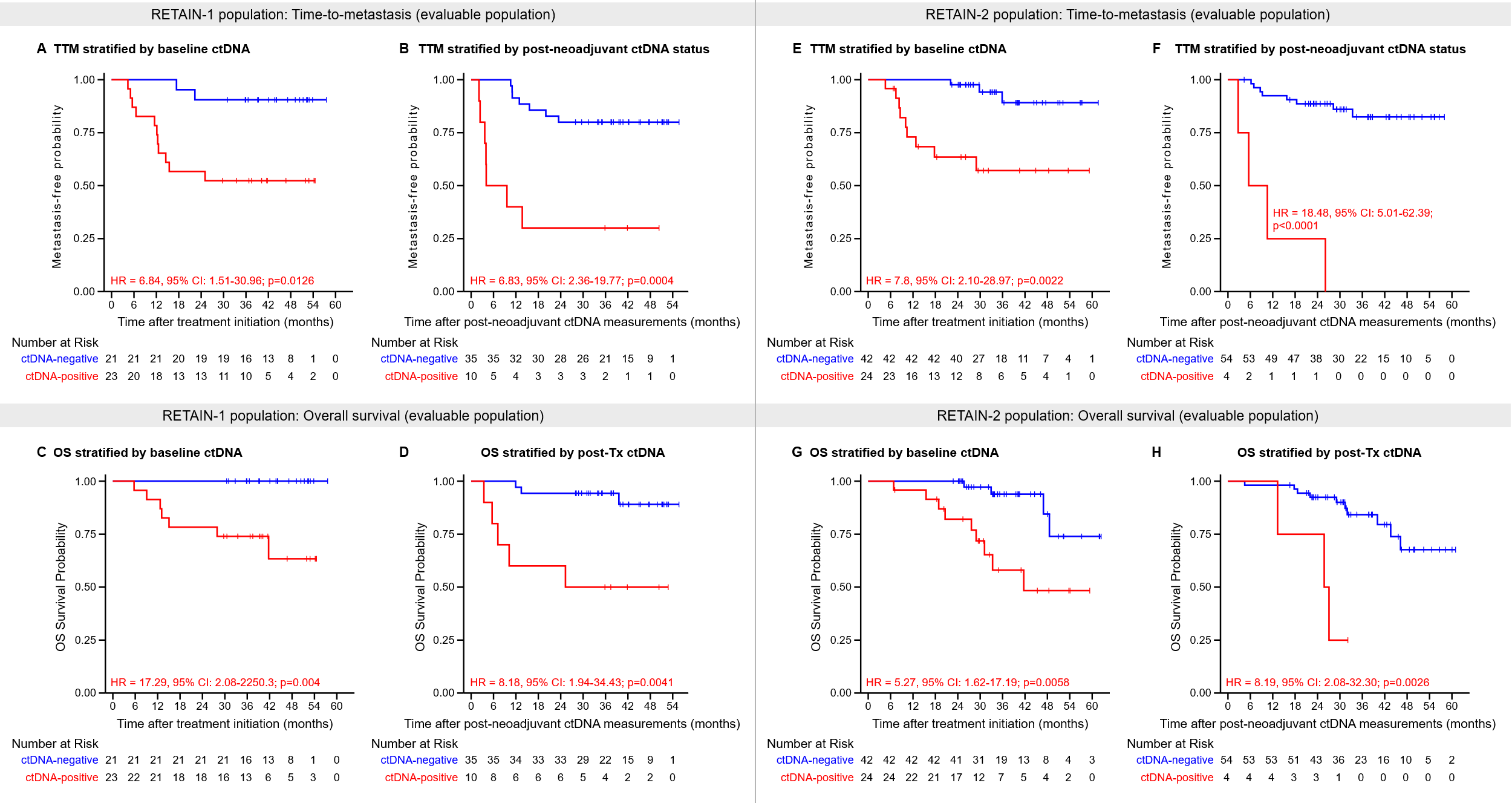

**Supplementary Figure 3: ctDNA-based prognostication stratified by clinical trial subgroup, RETAIN-1 (A-D) and RETAIN-2 (E-H)**

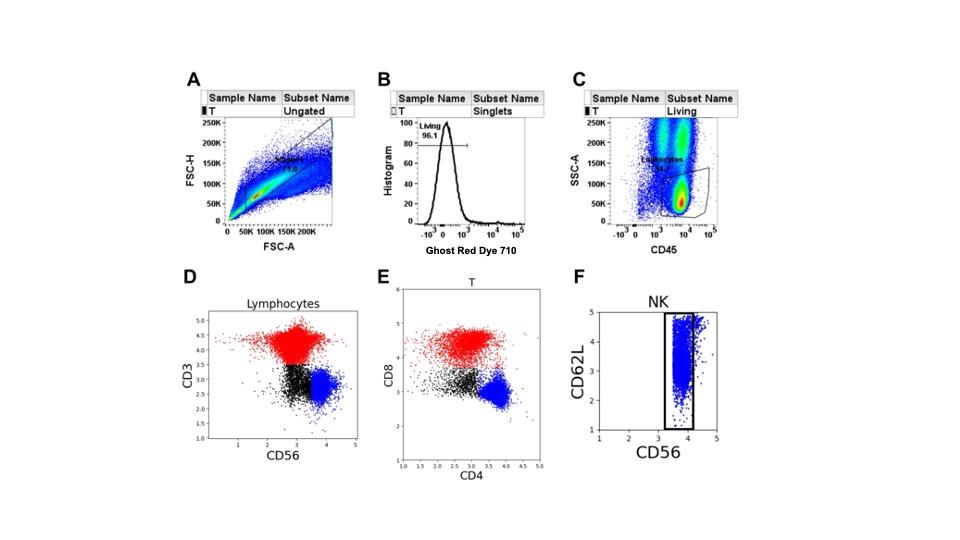

**Supplementary Figure 4: Gating strategy to define lymphocyte populations from a representative sample. (A)** Single cells were gated by forward scatter area and height. **(B)** Viable cells were gated as Ghost Red dye 710 negative. **(C)** Lymphocytes were selected as CD45^+^ side scatter low. Subsequent analysis of lymphocyte sub-populations was done in Python version 3.9 to facilitate mathematical analysis of the subsets. **(D)** T cells were identified as CD3^+^ and NK cells as CD3^-^ CD56^+^. **(E)** CD4^+^ and CD8^+^ T cells were selected with double positives included in the CD8^+^ population. **(F)** CD56^dim^ and CD56^bright^ subsets of NK cells were selected with reference to CD62L, which is highly expressed on the CD56^bright^ subset.

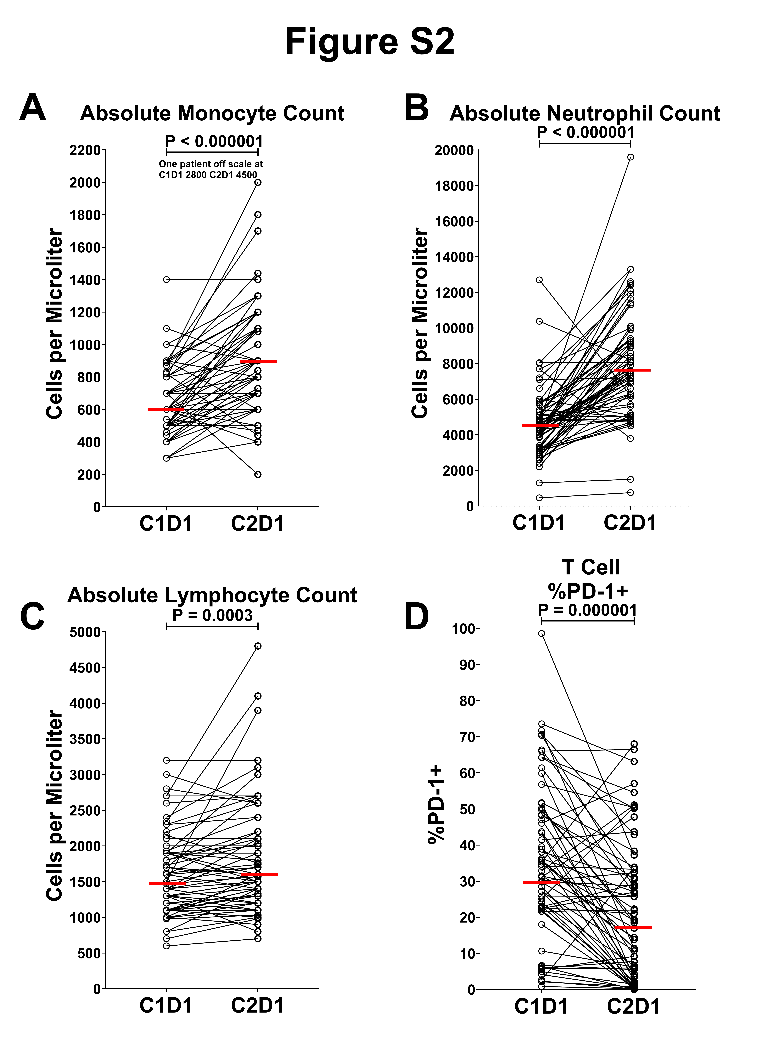

**Supplementary Figure 5: Changes in peripheral blood cell counts and T cell PD-1 expression from pre-treatment to cycle 2.** Pre-treatment (C1D1) and cycle 2 day 1 (C2D1) values for individual patients are shown as open circles connected by lines, with horizontal red bars indicating median values. P values were calculated using a Wilcoxon matched-pairs signed-rank test throughout. Absolute cell counts in panels **A–C** were derived from complete blood count (CBC) measurements. **(A)** Absolute monocyte counts at C1D1 and C2D1. One patient with an off-scale value at C1D1 (2,805 cells/µL) and C2D1 (4,586 cells/µL) is noted in the panel. **(B)** Absolute neutrophil counts at C1D1 and C2D1. **(C)** Absolute lymphocyte counts at C1D1 and C2D1. **(D)** %PD-1+ on CD3+ T cells gated as described above, at C1D1 and C2D1. Percent PD-1+ was calculated relative to a fluorescence-minus-one control tube containing all gating antibodies but no anti-PD-1 antibody.

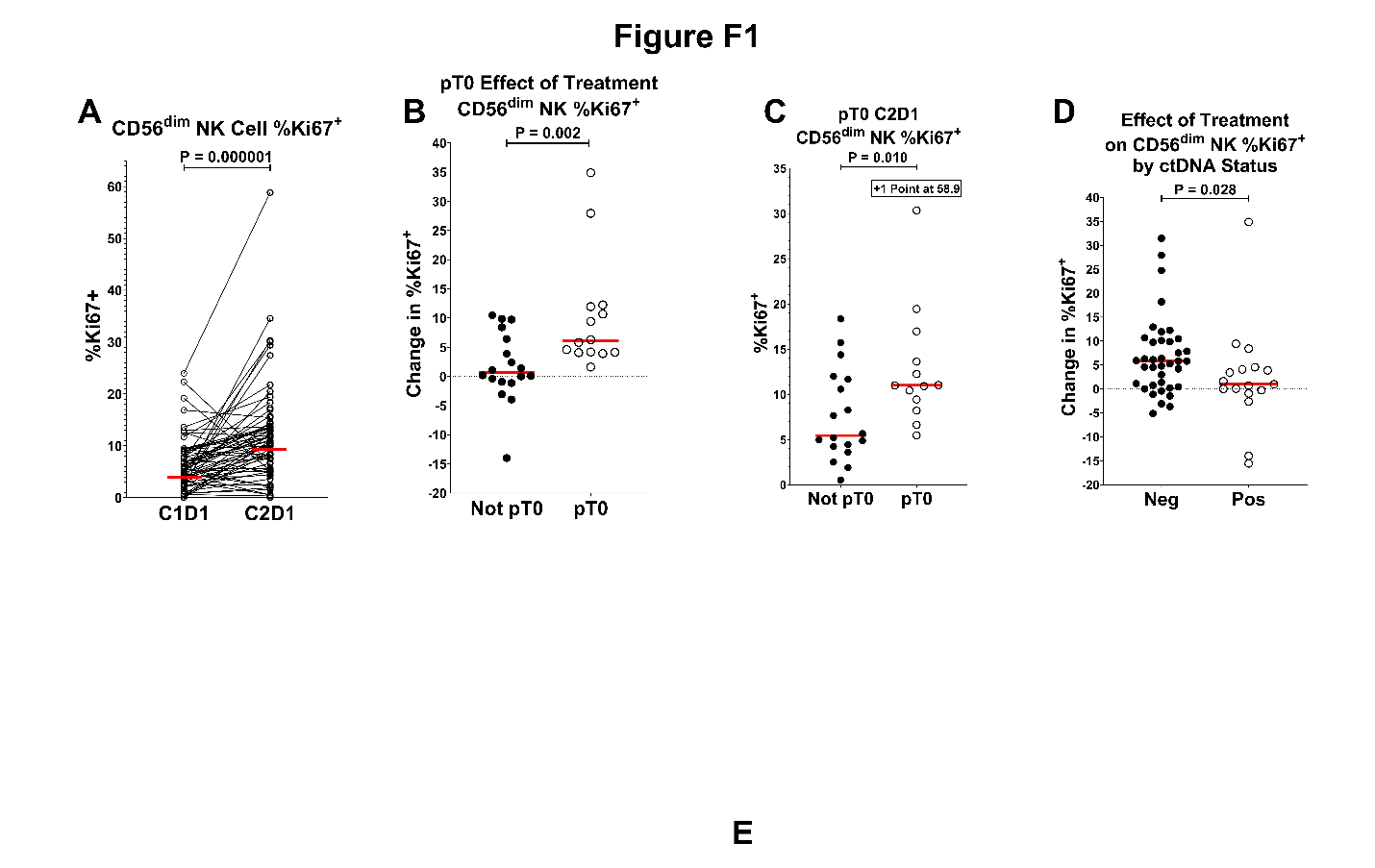

**Supplementary Figure 6. CD56dim NK cell Ki67 expression before and during treatment** CD56^dim^ NK cells were gated as viable singlet CD45^+^ CD3^-^ CD56^dim^ cells. Percent Ki67+ was measured relative to a fluorescence-minus-one control tube containing all gating antibodies but no Ki67 stain. **(A)** Pre-treatment (C1D1) and on-treatment (C2D1) %Ki67+ values for each patient in the entire cohort are shown, with individual patients connected by lines. Horizontal red bars indicate median values. Statistical comparison was performed using a Wilcoxon matched-pairs signed-rank test. For panels **(B)** and **(D)**, the effect of treatment was calculated as the change in %Ki67+ from pre-treatment (C1D1) to the beginning of cycle 2 (C2D1). **(B)** Change in %Ki67+ during treatment stratified by pathologic response at cystectomy, compared using a Wilcoxon rank-sum test. Patients with residual disease in the resected tissue (Not pT0) are shown as filled circles; patients with no residual disease (pT0) are shown as open circles. Horizontal red bars indicate median values. **(C)** %Ki67+ at C2D1 stratified by pathologic response at cystectomy. Groups and statistical test are as described in panel B. **(D)** Change in %Ki67+ on CD56^dim^ NK cells stratified by circulating tumor DNA (ctDNA) status. ctDNA-negative patients are shown as filled circles and ctDNA-positive patients as open circles. Statistical comparison was performed using a Wilcoxon rank-sum test. Horizontal red bars indicate median values throughout.

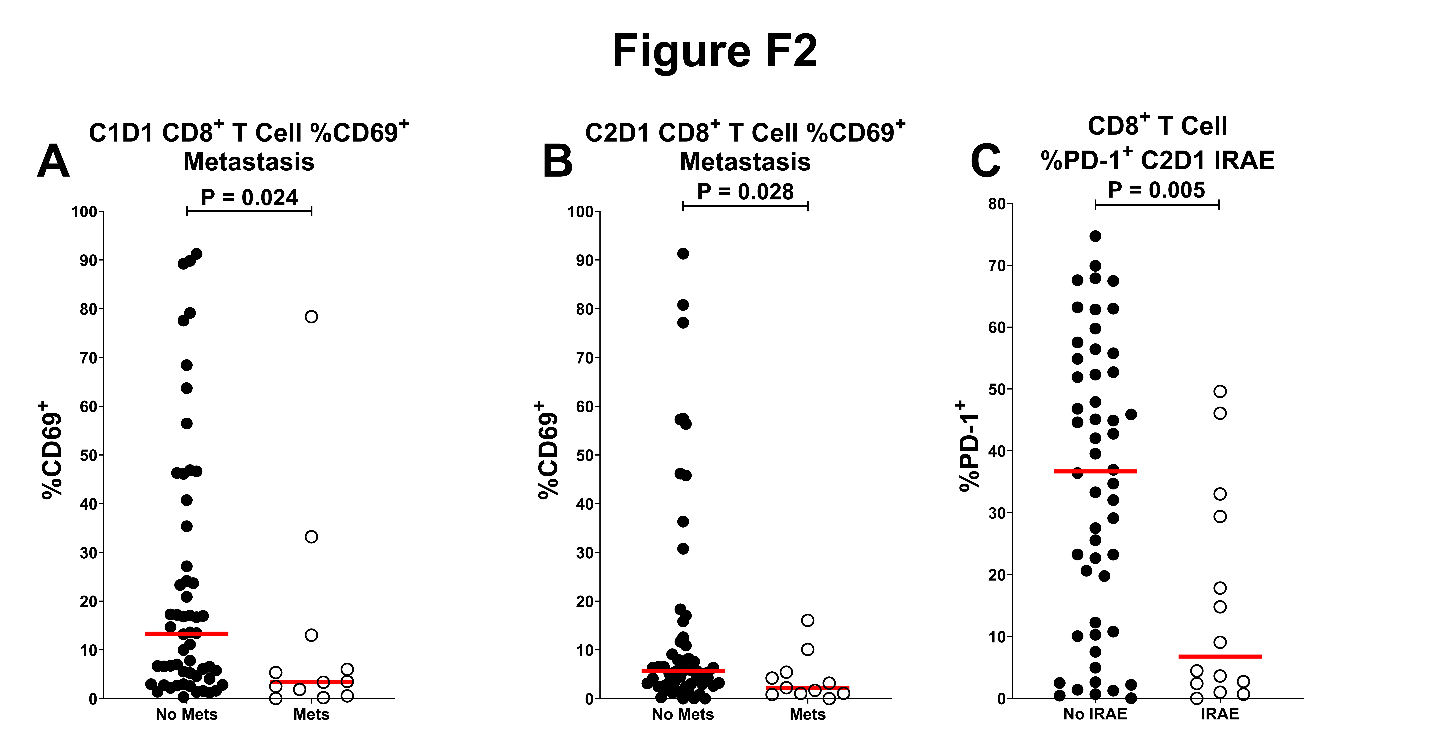

**Supplementary Figure 7.** **CD8^+^ T cell CD69 and PD-1 expression by metastatic status and immune-related adverse event (irAE) occurrence.** CD8^+^ T cells were gated as viable singlet CD45^+^CD3^+^CD8^+^ cells. Percent positive was measured relative to a fluorescence-minus-one control tube containing all gating antibodies but no CD69 or PD-1 staining antibody. Horizontal red bars indicate median values, and P values were calculated using a Wilcoxon rank-sum test throughout. **(A)** Pre-treatment (C1D1) %CD69^+^ on CD8^+^ T cells in patients without metastasis (filled circles) versus those with metastasis (open circles). **(B)** %CD69^+^ on CD8^+^ T cells measured at the start of cycle 2 (C2D1) in the same patient groups as in panel A. **(C)** %PD-1+ on CD8+ T cells at pre-treatment (C1D1) in patients who did not develop an irAE (filled circles) versus those who did (open circles).

| **Toxicity Type** | **Grade 3** | **Grade 4** | **Grade 5** |
| --- | --- | --- | --- |
| Thrombocytopenia | **6 (7.5%)** | **5 (6.2%)** | - |
| Mucositis | **8 (10.0%)** | - | - |
| Leukopenia | **6 (7.5%)** | **2 (2.5%)** | - |
| Anemia | **7 (8.8%)** | - | - |
| Acute kidney injury | **6 (7.6%)** | - | **1 (1.2%)** |
| Hyponatremia | **3 (3.8%)** | **1 (1.2%)** | - |
| Febrile neutropenia | **3 (3.8%)** | - | - |
| Pyelonephritis | **3 (3.8%)** | - | - |
| Pancytopenia | **2 (2.5%)** | - | **1 (1.2%)** |
| Aspartate aminotransferase increased | **2 (2.5%)** | **1 (1.2%)** | - |
| Dehydration | **2 (2.5%)** | - | - |
| Fatigue | **2 (2.5%)** | - | - |
| Blood bilirubin increased | **1 (1.2%)** | **1 (1.2%)** | - |
| Hypoxia | **1 (1.2%)** | **1 (1.2%)** | - |
| Neutrophil count decreased | - | **2 (2.5%)** | - |
| Urinary tract infection | **2 (2.5%)** | - | - |
| Syncope | **2 (2.5%)** | - | - |
| Diarrhea | **2 (2.5%)** | - | - |
| Fall | **2 (2.5%)** | - | - |
| Generalized muscle weakness | **2 (2.5%)** | - | - |
| Dysphagia | **1 (1.2%)** | - | - |
| Dizziness | **1 (1.2%)** | - | - |
| Alanine aminotransferase increased | **1 (1.2%)** | - | - |
| Enterocolitis | **1 (1.2%)** | - | - |
| Hypocalcemia | - | **1 (1.2%)** | - |
| Hyperglycemia | **1 (1.2%)** | - | - |
| Fever | **1 (1.2%)** | - | - |
| Insomnia | **1 (1.2%)** | - | - |
| Hypokalemia | **1 (1.2%)** | - | - |
| Hypomagnesemia | - | **1 (1.2%)** | - |
| Hypothyroidism | **1 (1.2%)** | - | - |
| Hyperkalemia | **1 (1.2%)** | - | - |
| Hyperlipidemia | **1 (1.2%)** | - | - |
| Hypoalbuminemia | **1 (1.2%)** | - | - |
| Multi-organ failure | - | - | **1 (1.2%)** |
| Nausea | **1 (1.2%)** | - | - |
| Lung infection | **1 (1.2%)** | - | - |
| Portal vein thrombosis | **1 (1.2%)** | - | - |
| Sepsis | **1 (1.2%)** | - | - |
| Urinary retention | **1 (1.2%)** | - | - |
| Thromboembolic event | **1 (1.2%)** | - | - |

^*^ All patients treated with at least one dose of AMVAC and nivolumab

**Supplementary Table S1: RETAIN-2 grade 3 to 5 Treatment-related Adverse Events of all treated patients** (including those unevaluable for the primary endpoint)^*^

| **Characteristic** | All ITT Patients (N=70) |
| --- | --- |
| Age |  |
| Median (range), y | 70.0 (47-83) |
| Sex, male, n (%) | 54 (77) |
| Race, n (%) |  |
| African American | 3 (4) |
| Asian | 2 (3) |
| White | 65 (93) |
| Baseline ECOG PS, n (%) |  |
| 0 | 58 (83) |
| 1 | 12 (17) |
| Clinical Stage |  |
| cT2 | 55 (79) |
| cT3 | 15 (21) |
| Additional Variant Histologic Features^a^ | 12 (17%) |
| AMVAC Cycles Received |  |
| 2 | 7 (10) |
| 3 | 63 (90) |

^a^ Squamous, adenocarcinoma, micropapillary, plasmacytoid

ECOG PS, Eastern Cooperative Oncology Group performance status; AMVAC, accelerated methotrexate, vinblastine, doxorubicin, cisplatin

**Supplementary Table S2**: Patient Demographics and Disease Characteristics at Baseline in RETAIN-1 ITT patients (N=70)

| **TTM** | | | | |
| --- | --- | --- | --- | --- |
| **Covariate** | **HR** | **HR lower 95% CI** | **HR upper 95% CI** | **p** |
| Post-NAT ctDNA positive vs negative | 3.92 | 1.63 | 9.22 | 0.0028 |
| Baseline ctDNA positive vs negative | 5.81 | 2.20 | 17.58 | 0.00028 |
| Post-NAT non-cCR vs cCR | 2.25 | 0.99 | 5.35 | 0.050 |

**Supplementary Table S3:** Multivariable Cox proportional hazards analysis of pre- and post-NAT ctDNA status and clinical CR status in overall ctDNA cohort

| **TTM** | | | | |
| --- | --- | --- | --- | --- |
|  | **ctDNA covariate** | | **Cohort covariate** | |
| **Covariate** | **HR (95% CI)** | **p** | **HR (95% CI)** | **p** |
| Baseline ctDNA status | 7.4 (2.74-19.96) | 7.7e-05 | 0.84 (0.38-1.85) | 0.67 |
| Post-NAT ctDNA status | 9.77 (4.22-22.57) | 9.6e-08 | 1.05 (0.46-2.38) | 0.13 |

| **OS** | | | | |
| --- | --- | --- | --- | --- |
|  | **ctDNA covariate** | | **Cohort covariate** | |
| **Covariate** | **HR (95% CI)** | **p** | **HR (95% CI)** | **p** |
| Baseline ctDNA status | 7.4 (2.46-22.33) | 0.00037 | 1.89 (0.75-4.78) | 0.18 |
| Post-NAT ctDNA status | 7.32 (2.84-18.81) | 3.58e-05 | 2.16 (0.84-5.56) | 0.11 |

**Supplementary Table S4:** Multivariable Cox proportional hazards analysis of baseline and post-neoadjuvant ctDNA status, including clinical trial cohort as a covariate

| **Web-link** | **Target Biomarker** | **Fluorophore** | **Clone** | **Catalog number** | **Manufacturer** |
| --- | --- | --- | --- | --- | --- |
| 1 | CD19 | BV605 | HIB19 | 302244 | BioLegend |
| 2 | CD3 | PE/Cy7 | SK7 | 344816 | BioLegend |
| 3 | CD38 | AF488 | HIT2 | 303512 | BioLegend |
| 4 | CD4 | Alexa Fluor 700 | OKT4 | 317426 | BioLegend |
| 5 | CD4 | PerCP/Cy5.5 | OKT4 | 317428 | BioLegend |
| 6 | CD8a | Pacific Blue | HIT8a | 300928 | BioLegend |
| 7 | CD45 | PE/Cy7 | 2D1 | 560178 | BD Pharmingen |
| 8 | CD56 | FITC | B159 | 562794 | BD Biosciences |
| 9 | CD56 | BUV563 | NCAM16.2 | 612928 | BD Horizon |
| 10 | CD62L | FITC | DREG-56 | 304838 | BioLegend |
| 11 | CD62L | PE | DREG-56 | 304806 | BioLegend |
| 12 | CD62L | BUV737 | SK11 | 749210 | BD OptiBuild |
| 13 | CX3CR1 | APC | 2A9-1 | 341608 | BioLegend |
| 14 | CD69 | APC | FN50 | 310910 | BioLegend |
| 15 | TIM3 | PerCP/Cy5.5 | F38-2E2 | 345016 | BioLegend |
| 16 | Viability Dye | Ghost Red 710 |  | 13-0871-T100 | Tonbo Biosciences |
| 17 | PD-1 | unlabeled | Pembrolizumab |  | Merck |
| 18 | Ki-67 | BUV395 | B56 | 564071 | BD Horizon |
| 19 | TOX | APC | REA473 | 130-118-335 | Miltenyi |
| 20 | TCF1 | Alexa Fluor 488 | C6309 | 6444S | Cell Signaling |
| 21 | Anti-human IgG4 (2° for PD-1) | PE | HP6025 | 9200-09 | SouthernBiotech |

**Supplementary Table S5 – Antibodies and reagents used for flow cytometry analyses.**

**Supplementary Methods and Results:**

**Immune Profiling by Flow Cytometry:**

To ensure consistency across participating sites, all samples were processed following overnight room-temperature storage in heparinized collection tubes. PBMC were isolated on Lymphoprep (Axis-shield POC AS, Oslo, Norway), per manufacturer’s instructions. The antibody panel (**Supplementary Table S5**) included markers defining T-cell and NK-cell subsets, activation (CD69), proliferation (Ki67), and exhaustion-associated states (Tox, PD-1, TCF1). Background gating controls were included for CD69, Ki67 and PD-1. Standardized gating strategies and compensation controls were applied. **(Supplementary Figure 4**) PD-1 expression was assessed using nivolumab to saturate surface PD-1 receptors followed by detection with an anti-human IgG4 secondary antibody.^26^ Intracellular stains for Tox, Ki67 and TCF1 were performed following fixation and permeabilization with FoxP3 Fixation/Permeabilization Buffer Set (eBiosciences/Invitrogen). Data were acquired on a 4-laser BD FACS Aria II flow cytometer and analyzed using Flowjo (version 10.8) and Python (version 3.9).

**Immunological correlates of clinical outcomes**

Peripheral immune correlates were analyzed in 68 evaluable RETAIN-2 patients with peripheral blood collected at baseline (C1D1) and after one treatment cycle (C2D1). Responders were defined as patients with ypT0 at cystectomy or, among those managed with active surveillance, chemoradiation, or intravesical therapy, no evidence of recurrence for ≥12 months.

Absolute leukocyte counts increased significantly during treatment, consistent with G-CSF effects, but were not associated with response (**Supplementary Figure 5**). The strongest immune associations with favorable outcomes involved proliferation of mature cytolytic CD56dim NK cells (**Supplementary Figure 6**). Ki67 expression on CD56dim NK cells increased significantly during treatment and was greater in patients achieving ypT0 and in patients who were ctDNA-negative at C2D1. Lower CD69 expression on CD8+ T cells was observed in patients who developed metastases, while lower PD-1 expression on CD8+ T cells during treatment was associated with immune related adverse events (**Supplementary Figure 7**).
